# DNA Aptamer Gold Nanoparticle Colorimetric Diagnostic Test Kit of Saliva Samples for SARS_Cov2 Virus Linked to Mobile Phone Application (Aptamex™)

**DOI:** 10.1101/2022.02.09.22269224

**Authors:** Siska Nurul Chotimah, Yudhistira Eka Putra, Jocelyn H. Ng, Agustyas Tijptaningrum, Nabilla Sonia Sahara, Evi Arfianti, Ridha Amaliah, Michael J. Edel, Mayreli Ortiz Rodriguez, Teresa Mairal Llerga, Ciara K. O’Sullivan, Steven Goh

**Affiliations:** Achiko AG, Tessinerplatz 7, CH-8002 Zurich, Switzerland; Faculty of Medicine, University of Lampung, Lampung 35141, Indonesia; Faculty of Medicine, University of Riau, Pekanbaru, Riau 28155, Indonesia; Madani Hospital, Biomolecular Laboratory, Pekanbaru, Riau 28155, Indonesia; Regenacellx.sl., Carrer de la Ciutat de Granada, 28, Barcelona 08005, Spain; Interfibio Research Group, Departament d’Enginyeria Quimica, Universitat Rovira i Virgili, Avinguda Països Catalans 26, 43007 Tarragona, Spain; Institució Catalana de Recerca i Estudis Avançats (ICREA), Passeig Lluís Companys 23, 08010 Barcelona, Spain

**Keywords:** SARS-CoV-2, virus, Spike protein, DNA aptamer, gold nanoparticles, colorimetric assay, test kit, diagnostic, clinical use

## Abstract

The development of the use of DNA aptamers for clinical applications to detect human diseases is at the forefront of research. Synthetic DNA aptamers are easy to generate, inexpensive, highly specific and have been postulated to replace antibodies for research and clinical use. Despite the considerable amount of published work on the use of DNA aptamers for research use, to date they have not been exploited for clinical diagnostics. SARS-CoV-2 virus is a pandemic causing a global disruptive event preventing people from travel, work and leisure activities resulting in a major health crisis, hospital overloads and a high death rate. The detection of SARS-CoV-2 virus in communities is therefore very important, especially for returning normality of life. The current gold standard for detection of SARS-CoV-2 virus is RT-PCR, a technique that is relatively expensive and most importantly with a slow turnaround time between sample procurement and result. This paper describes the development of a rapid, accurate, low-cost, facile to use assay for the detection of the SARS-CoV-2 spike protein in saliva. The assay exploits a simple system based on the use of a gold nanoparticle-aptamer complex, that can be easily produced and distributed, facilitating its deployment to the point-of-need, potentially reaching millions of individuals in resource-limited settings. The proposed colorimetric diagnostic test kit uses a SARS-CoV-2 DNA aptamer adsorbed on gold nanoparticles and salt-induced aggregation to detect the presence of SARS-CoV-2 Spike protein in saliva samples indicated by a color change of the gold absorbance spectrum that can be quantified by a spectrophotometer, linked to a mobile phone for data processing and analysis. The assay parameters were optimized and then tested in a field calibration clinical test in Indonesia. At a research level, a limit of detection of ca. 1.25 nM to synthetic spike protein (S1) was observed and a method to test human saliva samples developed. The DNA aptamer was specific to SARS-CoV-2, with minor cross-reactivity observed with MERS and SARS-CoV-1, but negligible cross-reactivity seen with common cold coronaviruses. A calibration clinical test of patients in Indonesia demonstrated a classification resulting in a > 97% sensitivity and a > 97% specificity compared with saliva RT-PCR test for SARS-CoV-2. Furthermore, the data indicates that anatomical location and sample type (swab vs saliva) can affect RT-PCR results. In conclusion, we have developed the use of a robust and reproducible aptamer-gold nanoparticle assay for clinical diagnostic use based on a colorimetric system that is cheap, simple, rapid, sensitive and can be employed for large scale testing of human populations for SARS-CoV-2 virus.

## Introduction

In late December 2019, a cluster of cases of pneumonia were reported in Wuhan, China, and a novel coronavirus was identified as the cause of the infection [1]. In January 2020 the genetic sequence of the virus was shared and the virus was named SARS-CoV-2 (Severe Acute Respiratory Syndrome Coronavirus 2). Likewise, the infection caused by this novel coronavirus was called COVID-19. On March 2020, the World Health Organization (WHO) announced COVID-19 outbreak as pandemic [2]. To date more than 300 million cases have been reported and the global death toll currently continues past 5 million [3].

SARS-CoV-2 is an enveloped RNA virus with a person-to-person transmission [4], that can cause a variety of symptoms such as fever, cough, sputum, myalgia, fatigue or diarrhea [5], as well as neurological symptoms [6]. Furthermore, the clinical course can differ from a mild disease to a severe pneumonia with acute respiratory distress syndrome [5]. The accurate detection of SARS-CoV-2 through respiratory sampling is critical for the prevention of further transmission and the timely initiation of treatment for COVID-19 [7]. The reported detection rate is different depending on the collection method and it is known that the naso-oropharyngeal detection rate varies from 25% to > 70% [8] and the saliva rate can range from 48% to > 90% [9].

Currently, the gold standard test to identify the virus is the quantitative Polymerase Chain Reaction (qPCR) on respiratory specimens collected mainly with a naso-oropharyngeal swab. However, it has some limitations as a screening tool, including the (i) need for trained personnel; (ii) requirement for typically laboratory-based equipment; (iii) long delay between sample procurement and assay result; and (iv) prohibitive expense as a screening tool for frequent use. We propose the use of a DNA aptamer-gold nanoparticle assay that detects the SARS-CoV-2 spike protein for a point-of-care (PoC) diagnostic system that is cheap, fast and specific as a solution for repeat screening of large populations for COVID-19. The proposed kit exploits the use of previously published aptamers for the SARS-CoV-2 spike proteins [10], using a well-established technique where the spectroscopic properties of gold nanoparticles [11,12] as they transition from disaggregated to aggregated states, can be used as a signal transduction mechanism for aptamer-based colorimetric [13] or lateral flow assays (LFA) [14]. Aptamers have been widely exploited as molecular recognition elements [15] in clinical diagnostics at a research level, as exemplified by the detection of the human epidermal growth factor receptor 2 for detection of cancer [16] dopamine in urine for Alzheimer’s disease [17], enteroviral nucleic acid sequences [18], platelet-derived growth factor-BB [19], as well as kanamycin [20].

qPCR is the gold standard for positive diagnosis of COVID-19, but this technique can even detect RNA even after the resolution of symptoms and the infectious phase of the virus, and a cycle threshold (C_T_) of 35 cycles is used for a positive diagnosis that is indicative of the ability of the virus to infect cells [21]. The C_T_ value has been used to predict the clinical course and mortality [22], suggesting a correlation between low C_T_ values and poorer prognostics and higher mortality [22-24].

## Methods

### Synthesis and characterization of gold nanoparticles

Gold Nanoparticles (AuNP) were made as previously published [25], based on the reduction of gold (III) chloride with sodium citrate. For the characterization of the AuNP, visible spectroscopy spectra were acquired on a Cary 100 Bio UV-Vis spectrometer (Varian)

### AuNP aptamer assay for synthetic S1 protein detection

To obtain the AuNP-aptamer complex, AuNP and the aptamer were incubated for 30 min at room temperature at the optimal concentrations obtained in a checkerboard assay, where the concentrations of aptamer and NaCl were optimised. Serial dilutions of SARS-CoV-2 spike protein were then added to the complex, followed by addition of the optimized concentration of NaCl. The spectra of the samples were recorded (300 – 700 nm) and the ratio of the absorbance of the aggregated particles (A640 nm) to the dispersed particles (A520 nm) was plotted against the concentration of the SARS-CoV-2 spike protein (S1). The limits of detection (LOD) were calculated, after interpolation in the calibration curve, as the absorbance of the blank plus three times the standard deviation of the blank (Ablank + 3 SDblank).

### Realtime RT-PCR for detection of SARS-CoV-2

Nasopharyngeal swab samples of the participants were lysed and RNA was extracted using a GeneRotex 96 and used to detect the presence of the SARS-CoV-2 virus using the PCR kit, Tianlong. Saliva samples were collected for RT-PCR according to instructions for the BioSaliva kit (Biofarma) or QuickSpit (Nalagenetics) where both require fasting from food and drink for at least 30 minutes. The RT-PCR was carried out according to manufacturer’s instructions using a Genteir 96E RT-PCR machine. The two target genes were ORF1ab, RdRp with ribonuclease P protein subunit P30 (RPP30) as the internal control (IC). The cutoff was at a Ct value of 32. Thus, all samples yielding results below or at a Ct value of 32 was considered a positive PCR result.

### Abbott Panbio rapid antigen test for detection of SARS CoV-2

Abbott Panbio rapid antigen tests were purchased and processed according to the manufacturer’s instructions. Briefly, all reagents were allowed to equilibrate at room temperature for 15 minutes, swabs were taken: swirled 5 times in both nostrils, the nasal mucous then added to buffer solution for 30 seconds, 5 drops added to the lateral flow assay and the test visually read after 15-20 minutes.

### Participants for Calibration test in patients

#### Work flow

Following patient information and consent to participate in the study, the patient is asked to fast from food and drink for at least 30 minutes. During the fasting period, nasopharyngeal and oropharyngeal (NPOP) swabs were taken. The NPOP samples are stored under 2-8°C until there were the minimum 16 samples required for RT-PCR.

Both NPOP and saliva PCR are carried out for fresh samples while frozen saliva samples would have had NPOP-PCR done previously (in August to October 2021) and will only be tested by saliva PCR in addition to AptameX.

After the NPOP swab, a second nasopharyngeal swab is taken and a rapid antigen test performed. When the patient has fasted for at least 30 minutes, the first saliva sample is collected according to the protocol of either QuickSpit or BioSaliva. Once the saliva sample is ready, the processed saliva sample is stored under 2-8°C until the PCR is carried out.

A second saliva sample is then collected according to the AptameX sampling protocol. The AptameX protocol does not require fasting. Nevertheless, it is interesting to note that where AptameX followed a fasting period, all saliva samples were clear and colorless, as required for the AptameX analysis.

#### Calibration tests used

AptameX™ saliva sample specimens were subjected to simultaneous testing. Simultaneous testing of AptameX™ kit on saliva, RT-PCR on same saliva and nasopharyngeal swab and rapid antigen test on nasopharyngeal swab sample was performed. All four tests were conducted within a 12-hour period to ensure the COVID status of a patient is consistent across all the tests:

1. NP/OP PCR (nasopharyngeal and oropharyngeal swab PCR)
2. Rapid Antigen Test (Abbott PanBio, nasopharyngeal swab sample)
3. Saliva PCR (BioSaliva or QuickSpit).
4. AptameX (saliva sample)

#### Patient Sample Specimens

Negative samples were obtained from people who have no symptoms and are in good health. They provided samples needed to run the 4 tests. Positive samples were obtained from frozen, COVID-positive saliva samples collected with RT-PCR status from nasopharyngeal swab recorded (C_T_ score recorded). The sample specimens were from symptomatic and asymptomatic subjects who were recruited from those who have had a PCR test with a Public Health Facility for contact tracing and agreed to AptameX™ saliva collection.

#### Final tests applied

The four tests described above (numbered 1 to 4) were applied to 31 fresh negative samples and two tests (numbered 3 and 4) applied to 30 frozen positive saliva samples with NP/OP RT-PCR covid status previous established.

#### Algorithm and statistics

The descriptive statistics including the frequencies, percentages with 95% confidence intervals, cross-tabulations, measures of sensitivity, specificity, positive and negative predictive values were calculated using the MS Excel and R software tool (R version 4.0.4 and R-Studio Version 1.4.1106). Data was acquired using a Metertech SP-880 Spectrophotometer, connected by Bluetooth using Capacitor v0.6.1 to an in-house developed application in Node.JS v12.19.0 and stored using a Postgres Database v12.7.

#### Negative Control in data analysis

Each result set was processed by first subtracting against a standardized negative control, and then secondly given the curve’s characteristics against a learnt secondary negative control, leaving a residual curve. This is then measured, and a score assigned. A result above the score is characterized as negative, and a score below is characterized as positive.

## Results

### The AptameX™ test kit

First, we developed a colorimetric assay using gold nanoparticles complexed with a DNA aptamer raised against the RBD region of the S1 region of the SARS-CoV-2 virus (Figure 1). Using a UV-VIS spectrophometer, saliva samples from patients or synthetic S1 protein is added to the gold nanoparticle-aptamer complex, where the presence of the S1 protein results in a quantifiable small shift in color from the red to blue spectrum (Figure 1A). The chemistry is based on the aggregation of gold nanoparticles as a result of the S1 viral protein induced displacement of the DNA aptamer (Figure 1B), with the aggregation being enhanced by the addition of NaCl (Figure 1B).

**Figure 1:**
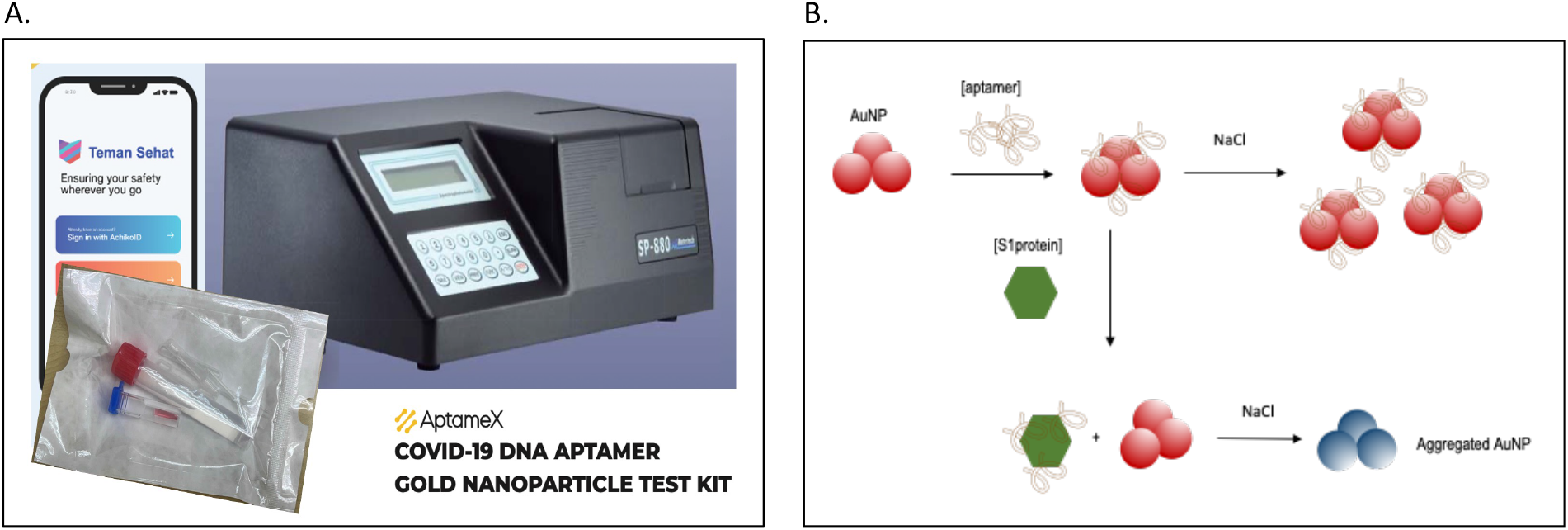
The AptameX™ test kit. **A**. The AptameX™ test kit: containing pre-filled 10-mL red cap tube for saliva sample, cuvette pre-filled with the AptameX™ reagent consisting of gold nanoparticles coated with a DNA aptamer specific for SARS-CoV-2 virus protein. SARS-CoV-2 virus protein binding to the DNA aptamer causes a color change of the conjugate that can be quantified by a Metertech SP-880 spectrophotometer. The result of the test kit is sent to a mobile phone using the Teman Sehat (TS: meaning “Health Buddy”) Mobile Phone Application. **B**. Schematic diagram of the assay concept, where DNA aptamers are adsorbed onto gold nanoparticles (AuNP). The SARS-CoV-2 spike protein binds to the adsorbed DNA aptamer, resulting in the aptamer being displaced from the Au-NP, resulting in AuNP aggregation and a shift in the visible absorption spectrum to longer wavelength, measured using a UV-VIS spectrophotometer.

### Limit of detection using synthetic S1 protein and specificity of DNA Aptamer

The limit of detection and specificity of the AptameX™ system was then evaluated using synthetic proteins of SARS-CoV-2 and other coronaviruses **(Figure 2)**. The data shows that the LOD is 1.25 nM and is highly specific for SARS-CoV-2 spike (S1) protein with a degree of cross-reactivity to the MERS spike protein (**Figure 2B**).

**Figure 2:**
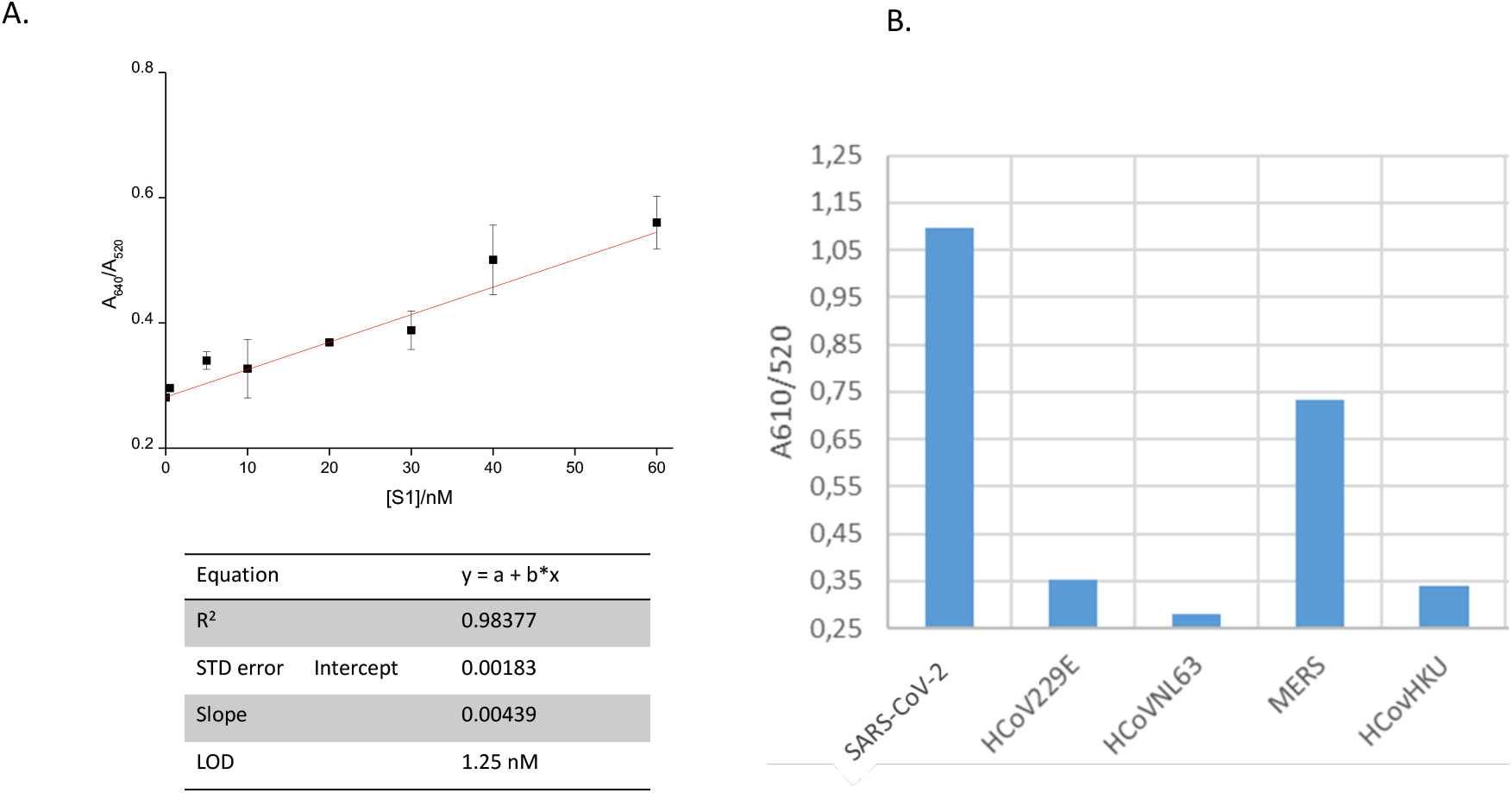
Limit of detection using synthetic S1 protein and specificity of DNA Aptamer. **A**. Graph showing the limit of detection of 1.25 nM of synthetic S1 protein. **B**. Graph demonstrating specificity of the DNA Aptamer tested on various coronavirus family members. **Note:** MERS is slightly detected.

### Clinical calibration test on patient saliva

Patient saliva samples were then tested. Results were obtained for 61 samples, 28 positive and 30 negative. 3 positive samples were discarded due to either contamination of the sample (1 sample), or a failure to sample sufficient quantities of saliva (2 samples). From the 58 samples, 30 gave a negative RT-PCR result (**Figure 3A**). Of the 28 RT-PCR positive individuals, AptameX™ classified 28 as positive (**Figure 3A**). All negatives were found negative (**Figure 3A**). The confusion matrix summarizing the results indicates no false negatives nor positives (**Figure 3C**). The overall result of the ROC analysis found AUC of 0.9988: > 97% sensitivity and a > 97% specificity (**Figure 3B**).

**Figure 3:**
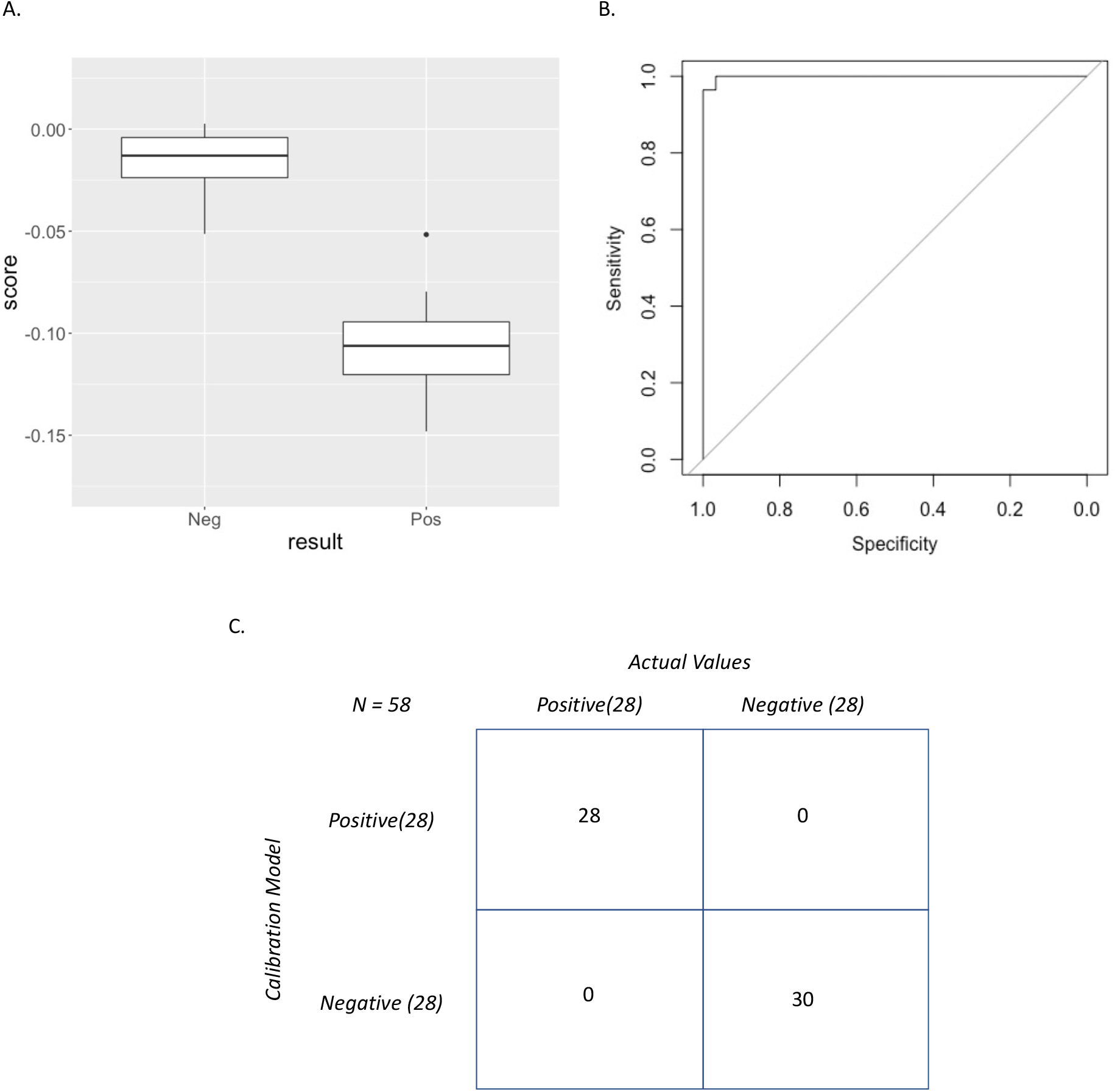
Clinical calibration test on patient saliva. **A**. Clustering analysis of positive and negative patient saliva test result from AptameX™. **B**. ROC analysis comparing AptameX™ (saliva sample) and RT-PRCR (saliva sample) result. **C.** Confusion matrix, classifying positive and negative results.

### Comparison of nasopharyngeal swab RTPCR vs Saliva RTPCR

We then investigated if the type of sample and location taken from influences the RTPCR result. We compared saliva sample to standard Swab methods **(Figure 4**). The results show that from the 61 samples tested (**Figure 4A**) that differences occur in the RT-PCR result depending on the anatomical location and sample type (**Figure 4B**). From a total of 30 positive samples obtained using the Swab methodology only 23 were found positive by saliva sampling methods, 18 with a C_T_ score above 25 and 5 samples with a C_T_ below 25 (**Figure 4B**).

**Figure 4:**
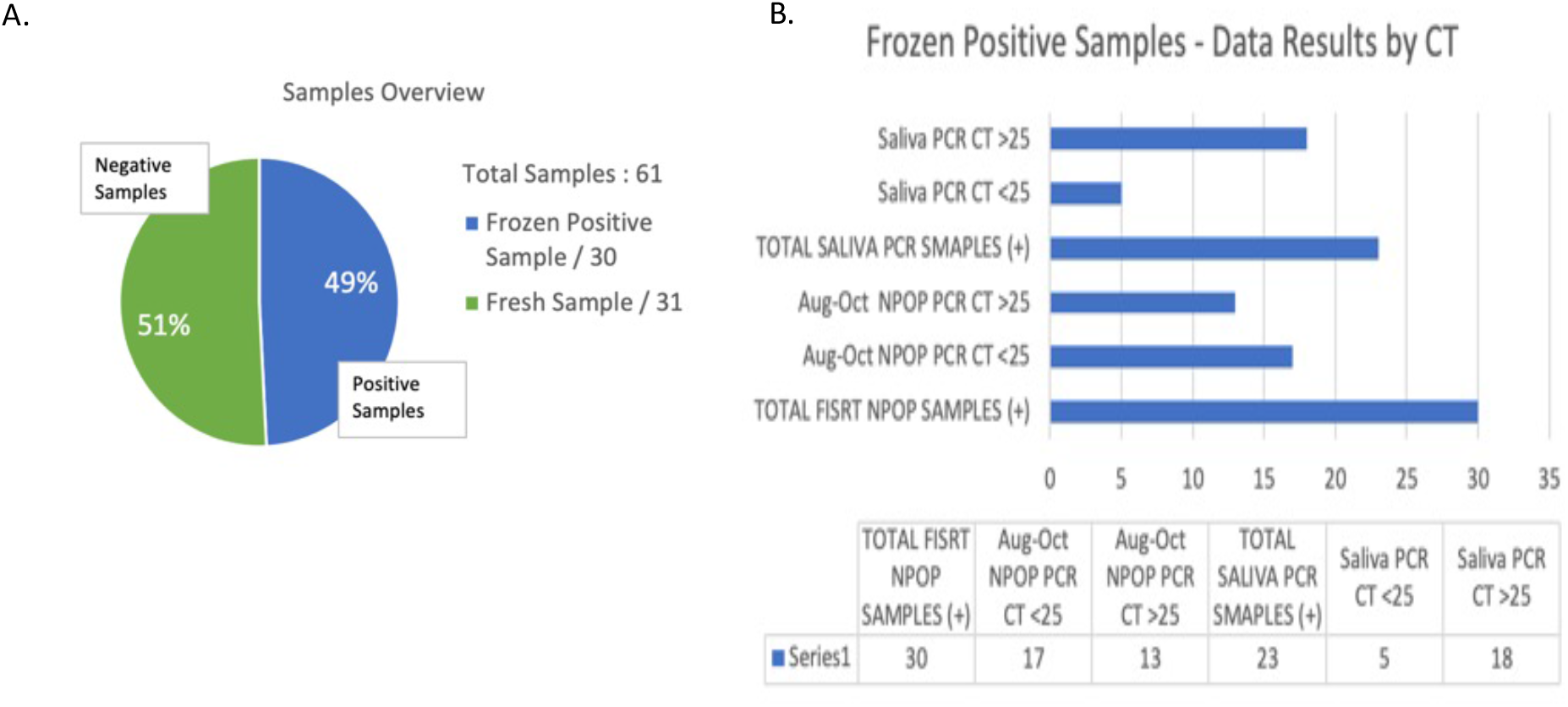
Comparison of nasopharyngeal swab RT-PCR vs saliva RT-PCR. **A**. Pie graph showing the number of patient samples used in the analysis. B. Best graph showing the number of positive cases from either swab or saliva methods based on the C_T_ score. **NPOP:** nasopharyngeal and oropharyngeal swab. The saliva PCR C_T_ values were higher, with the range of possible reasons including storage of the samples at -20 °C for more than a month, which may have resulted in degradation of the viral RNA.

## Discussion

Here we report the design and development of a test kit for the rapid detection of SARS-CoV-2 in patient saliva samples using a DNA aptamer-gold nanoparticle colorimetric test system. At a research level, a limit of detection of 1.25 nM to synthetic spike protein (S1) was observed, with negligible cross-reactivity to the common cold coronaviruses and a small degree of cross-reactivity with MERS. A calibration clinical test of patients in Indonesia demonstrated a classification resulting in a > 97% sensitivity and a > 97% specificity compared with a saliva RT-PCR test for SARS-CoV2.

The sensitivity of many antigen-based rapid diagnostic tests available in the market is comparable with RT-PCR at high viral loads (mean C_T_ value <25) but exhibit low sensitivity for low viral load specimens (mean C_T_ value >25). The trend is clear: as C_T_ values increase, these rapid test kits lose sensitivity. An official geometric mean (GM) C_T_ score of 28 could be considered an industry standard and the performance of all these currently available tests notably decrease from a C_T_ value of 28 [26-29]. The rationale for the use of rapid antigen tests is to be able to detect the SARS-CoV-2 virus in patients with high viral loads in order to identify and isolate them when they are contagious and thereby curb the pandemic. From a public health standpoint, in addition to accuracy, the other factor to consider with rapid tests is their frequent use. Even if all the cases can be correctly identified, there are still questions regarding the infectiousness such as degree and stage of disease, symptomology, and infection rates in the area of the patient concerned. Any test should ideally capture those that are at either side of the infection curve - those with early symptom on-set who will be infectious, and those in recovery who are still shedding virus [28-29]. However, to date, the duration of infectiousness is not fully understood [29]. A single test gives an answer at a single point in time and cannot be the basis of re-opening businesses and in controlling the pandemic, suggesting that testing should be frequent, carried out at least twice a week, in order to make these rapid tests effective in a pandemic [26-29], thus highlighting the critical requirement for a cheap, easy-to-use, rapid, robust assay, which can be facilely manufactured and deployed to all countries, independent of the local resource limitations. An important advantage of the AptameX™ assay is that it does not depend on components used in lateral flow based rapid antigen tests, such as membranes, conjugate pads, sample pads, plastic cassettes, which are now in huge demand, with the test simply requiring gold nanoparticles, DNA and NaCl, reagents which are widely available from a plethora of suppliers worldwide.

The performance of antigen-based rapid diagnostic tests varies greatly with clinical sensitivities ranging from 23% - 71% and 2-5 orders of magnitude less sensitive than RT-PCR [26-29]. Current rapid tests have high sensitivities at C_T_ values of 25 or lower, but at higher C_T_ values particularly above 28, performance declines [26-29].

Optimal C_T_ values should be lower than 25. A case in point: the example where Innova tests *“identified only two-thirds of the cases with C*_*T*_ *levels below 2*5” indicates that the laboratory which processed the samples performed at a level far from what should be required. Their C_T_ value of 25 would probably be equivalent to a C_T_ of 30 or higher if another lab would have processed the same samples [26-29]. The key message is that a third of the cases that were missed were probably infectious. If rapid tests should perform consistently at C_T_ values lower than 25, given the differences across laboratories, the C_T_ cut-off be so that diagnostic performance can be guaranteed. Mak et al. *Journal of Clinical Virology*, compared three different antigen tests and defined a “high” viral load at mean C_T_ value of 15.27-16.70 across the 3 commercially available rapid tests; “normal” at 23.21-24.77 and “low” at 31.83 [30]. Rapid tests then can be characterized across the three ranges of High, Normal and Low to understand its performance through its sensitivity curve. Thus, even if Ct values do not match across different laboratories, the trend will be apparent, and the test’s performance can be understood at either side of a given C_T_ value.

The different results between sampling methods or between testing laboratories could be caused by the quality of the RNA extracted from the sample, the anatomical location of sampling and/or type of sample (Figure 4). Viral load is considered equal or more in saliva compared to nasopharyngeal and oropharyngeal swab methods [27]. Because qPCR is a very sensitive method with an amplification step it suggests that RNA quality could be an important factor in determining if a patient is positive or negative for SARS-CoV-2 and to date is not part of quality control in the standard operating procedure for testing platforms.

In conclusion, AptameX™ Covid-19 test kit we report that AptameX™ performs with a > 97% sensitivity and a > 97% specificity. A limit of detection of 1.25 nM to synthetic spike protein (S1) was observed and specific to SARS-CoV-2, with minor cross-reactivity observed with MERS and SARS-CoV-1, but negligible cross-reactivity seen with common cold coronaviruses. AptameX™ has a low-price point, and exhibits comparable or better sensitivity to existing rapid tests. AptameX™ aims to be the solution in frequent mass-testing and screening.

## Data Availability

All data produced in the present study are available upon reasonable request to the authors

## Declaration

The research in this manuscript is original and not submitted nor published elsewhere and all authors agree to publish this work. All ethics, patient information and permissions for clinical testing obtained from Indonesian authorities.

## Acknowledgments

The authors would like to acknowledge Ace Goh, Windiaprana Ramelan, and Oktanita for technical assistance, management and product packaging, and to all the laboratory staff of RSUD Arifin Achmad, Laboratory of LONTAR Faculty of Medicine Universitas Riau and Official Health Department Pekanbaru City, Indonesia.

## References

[1] Wang, C., et al., A novel coronavirus outbreak of global health concern. Lancet, 2020. 395(10223): p. 470–473.

[2] The WHO timeline COVID-19, https://www.who.int/news-room/detail/27-04-2020-who-timeline---covid-19.

[3] John Hopkins Coronavirus Resource Center, https://coronavirus.jhu.edu/map.html

[4] Li Q, Guan X, Wu P, et al. Early transmission dynamics in Wuhan, China, of novel coronavirus-infected pneumonia. N Engl J Med 2020; published online Jan 29. DOI:10.1056/NEJMoa2001316.

[5] Fei Zhou et al., Lancet 2020; 395: 1054–62, March 9, 2020, https://doi.org/10.1016

[6] Abigail Whittaker et al., Neurological Manifestations of COVID-19: A systematic review and current updatem Acta Neurol Scand. 2020 Jun 2: doi: 10.1111/ane.13266

[7] Mohammadi A, Esmaeilzadeh E, Li Y, Bosch RJ, Li JZ. SARS-CoV-2 detection indifferent respiratory sites: A systematic review and meta-analysis. EBioMedicine. 2020 Sep;59:102903.

[8] Yu F, Yan L, Wang N, et al. Quantitative detection and viral load analysis of SARS-CoV-2 in infected patients. Clin Infect Dis 2020.

[9] Wu J, Liu J, Li S, et al. Detection and analysis of nucleic acid in various biological samples of COVID-19 patients. Travel Med Infect Dis 2020:101673.

[10] Song, Y.; Song, J.; Wei, X.; Huang, M.; Sun, M.; Zhu, L.; Lin, B.; Shen, H.; Zhu, Z.; Yang, C. Discovery of aptamers targeting the receptor-binding domain of the SARS-CoV-2 spike glycoprotein. Anal. Chem. 2020, 92, 9895–9900.

[11] W. Zhou, X. Gao, D. B. Liu and X. Y. Chen, Chemical Reviews, 2015, 115, 10575–10636.

[12] X. M. Ma, M. Sun, Y. Lin, Y. J. Liu, F. Luo, L. H. Guo, B. Qiu, Z. Y. Lin and G. N. Chen, Chinese, Journal of Analytical Chemistry, 2018, 46, 1–10.

[13] J. H. Soh, Y. Y. Lin, S. Rana, J. Y. Ying and M. M. Stevens, Analytical Chemistry, 2015, 87, 7644–7652.

[14] W. L. Zhou, W. J. Kong, X. W. Dou, M. Zhao, Z. Ouyang and M. H. Yang, Journal of Chromatography B-Analytical Technologies in the Biomedical and Life Sciences, 2016, 1022, 102–108.

[15] Reid R, Chatterjee B, Das SJ, Ghosh S, Sharma TK. Application of aptamers as molecular recognition elements in lateral flow assays. Anal Biochem. 2020 Mar 15;593:113574. Review.

[16] Ranganathan V, Srinivasan S, Singh A, DeRosa MC. An aptamer-based colorimetric lateral flow assay for the detection of human epidermal growth factor receptor 2 (HER2). Anal Biochem. 2020 Jan 1;588:113471.

[17] Dalirirad S, Steckl AJ. Lateral flow assay using aptamer-based sensing for on-site detection of dopamine in urine. Anal Biochem. 2020 May 1;596:113637.

[18] Veeren Chauhan Mohamed M Elsutohy C Patrick McClure Will Irving Neil Roddis Jonathan W Aylott. Gold-Aptamer-Nanoconstructs Engineered to Detect Conserved Enteroviral Nucleic Acid Sequences posted on 24.06.2019. Chem Rxiv.

[19] Razmi N, Baradaran B, Hejazi M, Hasanzadeh M, Mosafer J, Mokhtarzadeh A, de la Guardia M. Recent advances on aptamer-based biosensors to detection of platelet-derived growth factor. Biosens Bioelectron. 2018 Aug 15;113:58–71.

[20] Ou Y, Jin X, Liu J, Tian Y, Zhou N. Visual detection of kanamycin with DNA-functionalized gold nanoparticles probe in aptamer-based strip biosensor. Anal Biochem. 2019 Dec 15;587:113432.

[21] Bullard J, Dust K, Funk D, Strong JE, Alexander D, Garnett L, Boodman C, Bello A, Hedley A, Schiffman Z, Doan K, Bastien N, Li Y, Van Caeseele PG, Poliquin G. Predicting Infectious Severe Acute Respiratory Syndrome Coronavirus 2 From Diagnostic Samples. Clin Infect Dis. 2020 Dec 17;71(10):2663.

[22] Rao SN, Manissero D, Steele VR, Pareja J. A Systematic Review of the Clinical Utility of Cycle Threshold Values in the Context of COVID-19. Infect Dis Ther. 2020 Sep;9(3):573–586.

[23] Huang JTR, Ran RX, Lv ZH, et al. Chronological changes of viral shedding in adult inpatients with COVID-19 in Wuhan, China. Clin Infect Dis. 2020; 631.

[24] Zheng SF, Fan J, Yu F, et al. Viral load dynamics and disease severity in patients infected with SARS-CoV2 in Zhejiang province, China, January-March 2020: retrospective cohort study. BMJ. 2020;369.

[25] Lerga TM, Skouridou V, Bermudo MC, Bashammakh AS, El-Shahawi MS, Alyoubi AO, O’Sullivan CK. Gold nanoparticle aptamer assay for the determination of histamine in foodstuffs. Mikrochim Acta. 2020 Jul 16;187(8):452.

[26] Chandima Jeewandara, Dinuka Guruge, Pradeep Pushpakumara, et. al. Sensitivity and Specificity of Two WHO Approved SARS-CoV2 Antigen Assays in Detecting Patients with SARS-CoV2 Infection. BMC Infectious Diseases. Under review 23-09-2021. DOI: 10.21203/rs.3.rs-618824/v1

[27] Anne L. Wyllie, John Fournier, Arnau Casanovas-Massana, et. al. Saliva is more sensitive for SARS-CoV-2 detection in COVID-19 patients than nasopharyngeal swabs. medRxiv 2020.04.16. 20067835; doi: https://doi.org/10.1101/2020.04.16.20067835

[28] Singanayagam A, Patel M, Charlett A, Lopez Bernal J, Saliba V, Ellis J, Ladhani S, Zambon M, Gopal R. Duration of infectiousness and correlation with RT-PCR cycle threshold values in cases of COVID-19, England, January to May 2020. Euro Surveill. 2020 Aug;25(32):2001483.

[29] Peeling RW, Heymann DL, Teo YY, Garcia PJ. Diagnostics for COVID-19: moving from pandemic response to control. Lancet. 2021 Dec 20: S0140-6736(21)02346-1.

[30] Mak GC, Cheng PK, Lau SS, Wong KK, Lau CS, Lam ET, Chan RC, Tsang DN. Evaluation of rapid antigen test for detection of SARS-CoV-2 virus. J Clin Virol. 2020 Aug;129: 104500. doi: 10.1016/j.jcv.2020.104500.

